# The change pattern and significance of IgM and IgG in the progress of COVID-19 disease

**DOI:** 10.1101/2020.07.20.20157446

**Authors:** Xuzhen Qin, Jun Shen, Erhei Dai, Haolong Li, Guodong Tang, Lixia Zhang, Xin Hou, Minya Lu, Xian Wu, Simeng Duan, Jingjia Zhang, Yongzhe Li

## Abstract

**Background:** To investigate the significance of IgM and IgG in the progress of COVID-19.

**Method:** A multicenter cross-sectional study conducted in suspected and confirmed patients from four hospitals of China and a cohort study to identify the change pattern and significance in the process of COVID-19 disease.

**Results:** A total of 571 patients were enrolled in the cross-sectional study, including 235 confirmed SARS-CoV-2 infection with 91.9% patients IgG positive and 92.3% IgM positive. 30 patients diagnosed with SARS-CoV-2 infection were enrolled in the cohort study for flowing-up in 20 days. The peak of IgM and IgG reached in 10^th^ and 20 ^th^ day separately after symptom onset. The relationship between clinical classification and serological antibodies were analysed. The positive rate of COVID-19 IgG and IgM increased along with the clinical classification and the delay of treatment time.

**Conclusion:** We demonstrated the kinetics of IgM and IgG SARS-CoV-2 antibody in COVID-19 patients, which may contribute to explain the results of IgM and IgG SARS-CoV-2 antibody test and predict the prognosis of COVID-19.

## Introduction

2019 coronavirus disease (COVID-19) caused a great pandemic in all of the world. Until June 1, more than 6 million cases have been diagnosed COVID-19 and the mortality reached nearly 0.6% according to World Health Organization (WHO) reports. Chinese government and medical workers have taken great effort to prevent the distribution of this pandemic in nationwide. Facing with this unknown infectious disease, how to treat and avoid virus transmission is an urging question. The Chinese diagnosis and therapeutic guideline has been published 7 times[1]. More and more detailed information of COVID-19 got to known by the public.

One of the dilemma in the treatment of COVID-19 is the false negative rate of nucleic acid tests. The reasons cover several aspects including the low virus concentration in the upper respiratory tract, unstandardized sample collecting method, various performance of gene application method, decrease of viral load after one-week since disease onset[2,3]. Antibody tests has been confirmed a good supplement for nucleic acid tests. Since the immunity reaction were involved in the progress of COVID-19 disease, serological assays to detect antibodies has been developed and practiced in many countries [4]. It was used as an immunity passport or proof for the previous infection or asymptomatic infection or immunization. Yet, there’s still many challenges and unknown knowledge for the clinical practice of COVID-19 antibodies, including the stable performance of the products, the prevalence of antibodies in various region, the protection role of antibodies and the duration and relationship with the clinical stage. To better understand these questions with COVID-19 serological tests, we arrange this study in multi-center of China.

## Methods

### Patients

This study is a multi-center study with patients from Peking Union Medical College Hospital, Tianjin Haihe Hospital, the Fifth Hospital of Shijiazhuang and Zhongnan Hospital of Wuhan University. It divided into a cross-sectional study with 576 patients and a cohort study with 30 patients. All the clinical data were retrieved from Laboratory Information System (LIS) and Hospital Information System (HIS) of enrolled hospitals. This study has been approved by the ethnic committee of Peking Union Medical College Hospital. Informed consent is waived because of the use of the remaining samples.

### Measurement and classification

IgM and IgG antibody against COVID-19 in plasma and serum were tested by a chemilumin-escence assay or an immuno-chromatographic assay developed by Beier bioengineering company. Cutoff values of IgM and IgG were both used as 5 CU. Clinical classification referred to Chinese Recommendations for Diagnosis and Treatment of SARS-CoV-2 infection version 7 as light, normal, heavy, critical. According to the time interval from sample collecting to disease onset, patients were divided into three groups as ≤7days, 8-15days and >15days.

### Statistical Analysis

Continue variables were described by mean with standard deviation for normal distribution data and median with interquartile range (IQR) for skewed distribution data. Comparison between groups using Kruskal-Wallis tests for continuous variables and chi-square test for categorical variables. All statistical analysis was conducted by SPSS 12.0 (SAS Institute, Cary, NC, USA).

## Results

### The demographic characters of enrolled patients

A total of 571 patients were enrolled in this study, of which 144 patients from Beijing, 147 patients from Tianjin, 29 patients from Wuhan, and 241 from Shijiazhuang, respectively. 235 patients were diagnosed COVID-19 disease. The basic demographic is shown in Table 1. Among of enrolled patients, 235 participants were confirmed COVID-19 and 336 participants excluded COVID-19 according to the clinical diagnosis. There were little more male patients than female patients in this study. COVID-19 positive patients in Wuhan and COVID-19 negative patients in Tianjin were older than other patients. In 235 confirmed COVID-19 patients, altogether 216 of 240 (91.9%) patients tested IgG positive and 217 of 240 (92.3%) tested IgM positive.

**Table 1.**
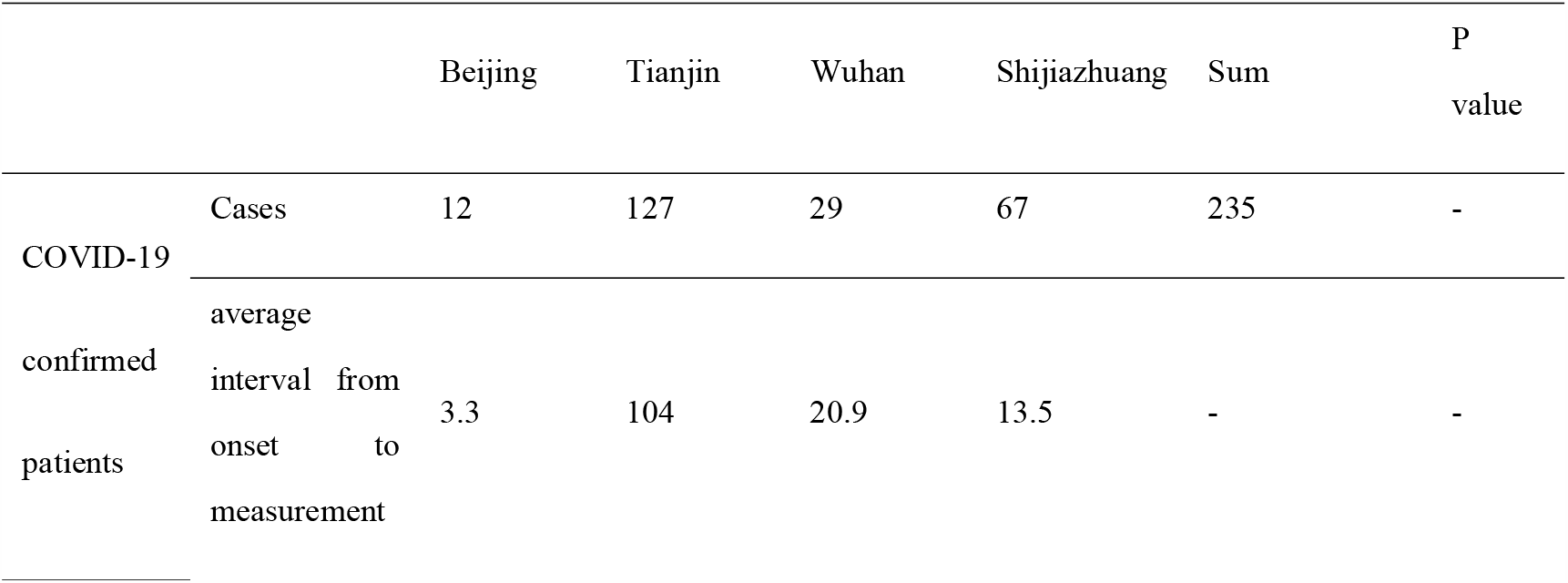
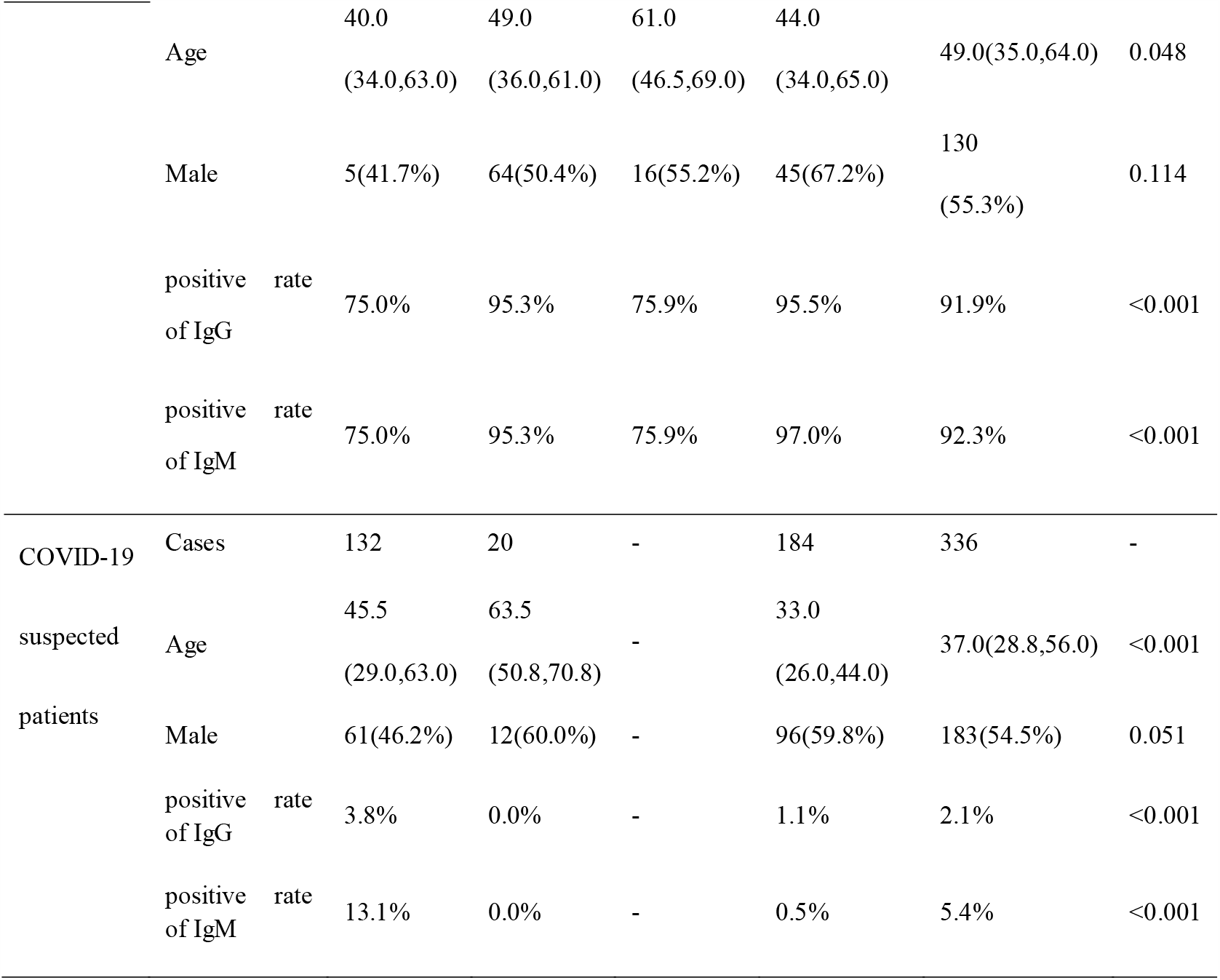
The demographic of enrolled patients in cross-sectional study.

### Antibodies concentration stratified by clinical classification

41 patients diagnosed COVID-19 with clinical classification information from Beijing and Wuhan were analyzed in Table 2. Although the number of patients with light symptom was only one, we could see the increase of median age, the concentration and positive rate of COVID-19 IgG and IgM along with the aggravation of illness.

**Table 2.** Antibodies concentration stratified by clinical classification.

|  | Clinical Classification |  |  | Sum | P<br>value |
| --- | --- | --- | --- | --- | --- |
|  | Light | Normal | Severe |  |  |
| N | 1 | 34 | 6 | 41 | - |
| Age | 25 | 52.5(39.8, 64.8) | 67.0(55.5,71.5) | 60.0(39.5,67.5<br>) | 0.09<br>1 |
| Male | 0.0% | 50.0% | 33.3% | 51.2% | 0.36<br>6 |
| Concentration<br>of | 9.1 | 22.2 (3.9, 89.4) | 157.9(28.8,190.7 | 30.6(8.4, | 0.05 |
| COVID-19 IgG |  |  |  | 129.4) | 6 |
| Positive rate of COVID-19 IgG | 100.0% | 70.6% | 100.0% | 75.6% | 0.25<br>6 |
| Concentration of COVID-19 IgM | 8.17 | 27.9 (4.4, 145.6) | 29.7 (8.6, 188.6) | 25.5(5.2,152.7 ) | 0.67<br>7 |
| Positive rate of COVID-19 IgM | 100.0% | 73.5% | 83.3% | 75.6% | 0.74<br>2 |

### Comparison of COVID-19 IgG and IgM in diagnosed patients between regions and symptom onset intervals

All patients diagnosed in cross-sectional study has collected the time interval from symptom onset to sampling. COVID-19 IgG and IgM positive rate stratified by symptom onset interval was shown in Figure 1A. The positive rate gradually increased with the delay of treatment time. Longer than 15 days the positive rate of COVID-19 IgG was closed to 100%. Figure 1B and 1C display the median of concentration of COVID-19 IgG and IgM in various regions. Obviously, the concentration of COVID-19 IgG and IgM rose up in 8-15 days in all regions. The concentration of IgM in Wuhan and IgG in Tianjin ascended distinctively step by step along with time interval.

**Figure 1.**
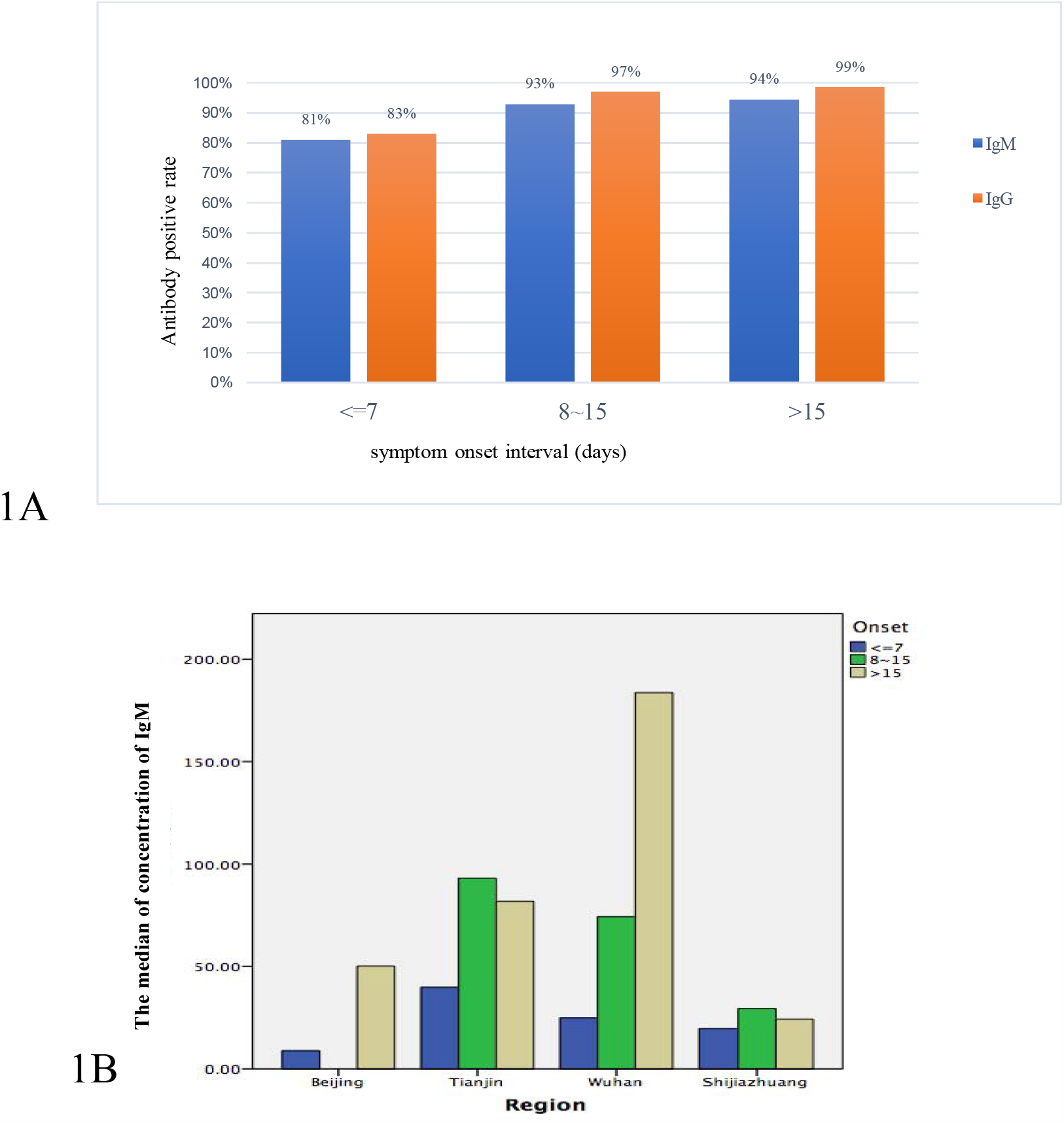
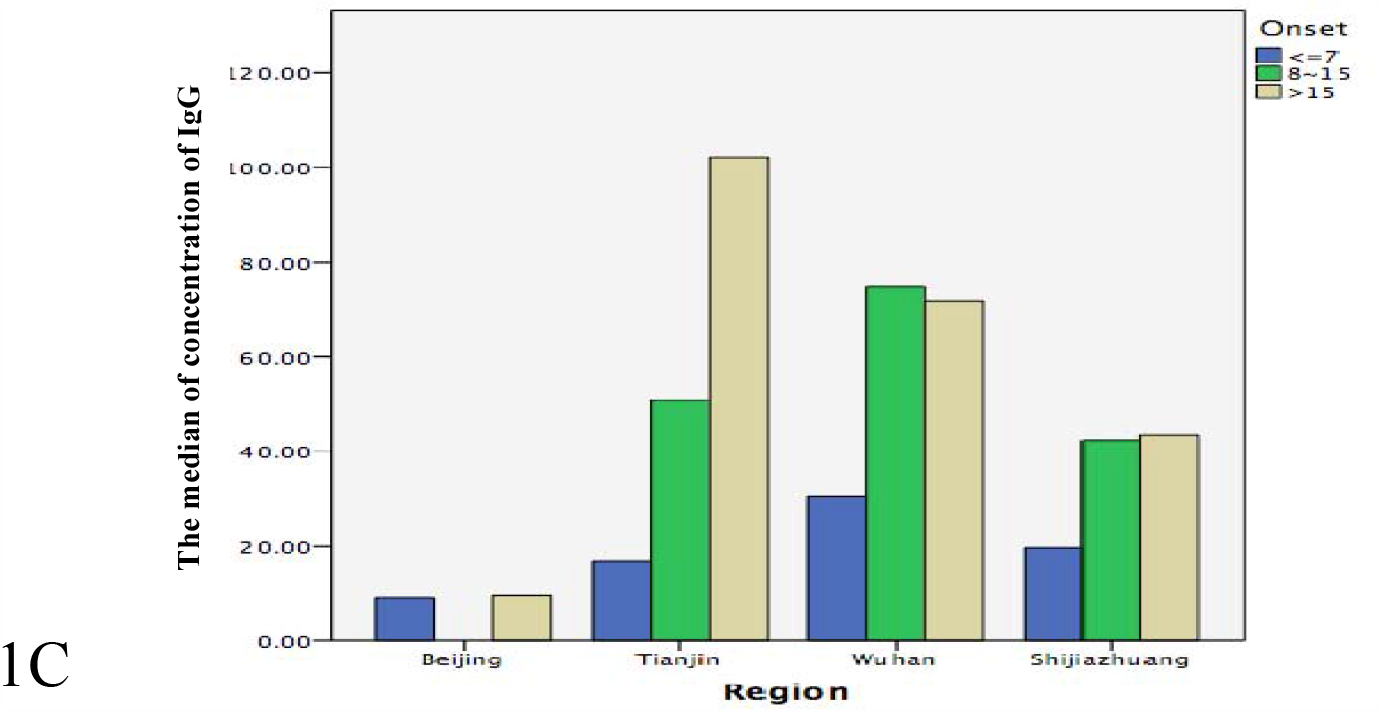
The positive rate and concentration comparison of COVID-19 IgG and IgM in patients diagnosed stratified by regions and symptom onset interval. 1A The positive rate of COVID-19 IgG and IgM stratified by symptom onset interval 1B The positive rate of COVID-19 IgM stratified by symptom onset interval and region 1C The positive rate of COVID-19 IgG stratified by symptom onset interval and region

### The dynamic characteristics of IgM and IgG in the cohort study

30 patients diagnosed with COVID-19 disease from Tianjin were enrolled in this cohort study to investigate the dynamic change of IgM and IgG concentration. The change pattern of IgM and IgG are shown in Figure 2. The concentration of IgM was higher than IgG before the 15^th^ day after the symptom onset. Subsequently, the concentration of IgG was higher than IgM. On the 10^th^ day the concentration of IgM reached the peak after the symptom onset and then decreased slowly. While the concentration of IgG on the 20 ^th^ day still kept the tendency of increase.

**Figure 2.**
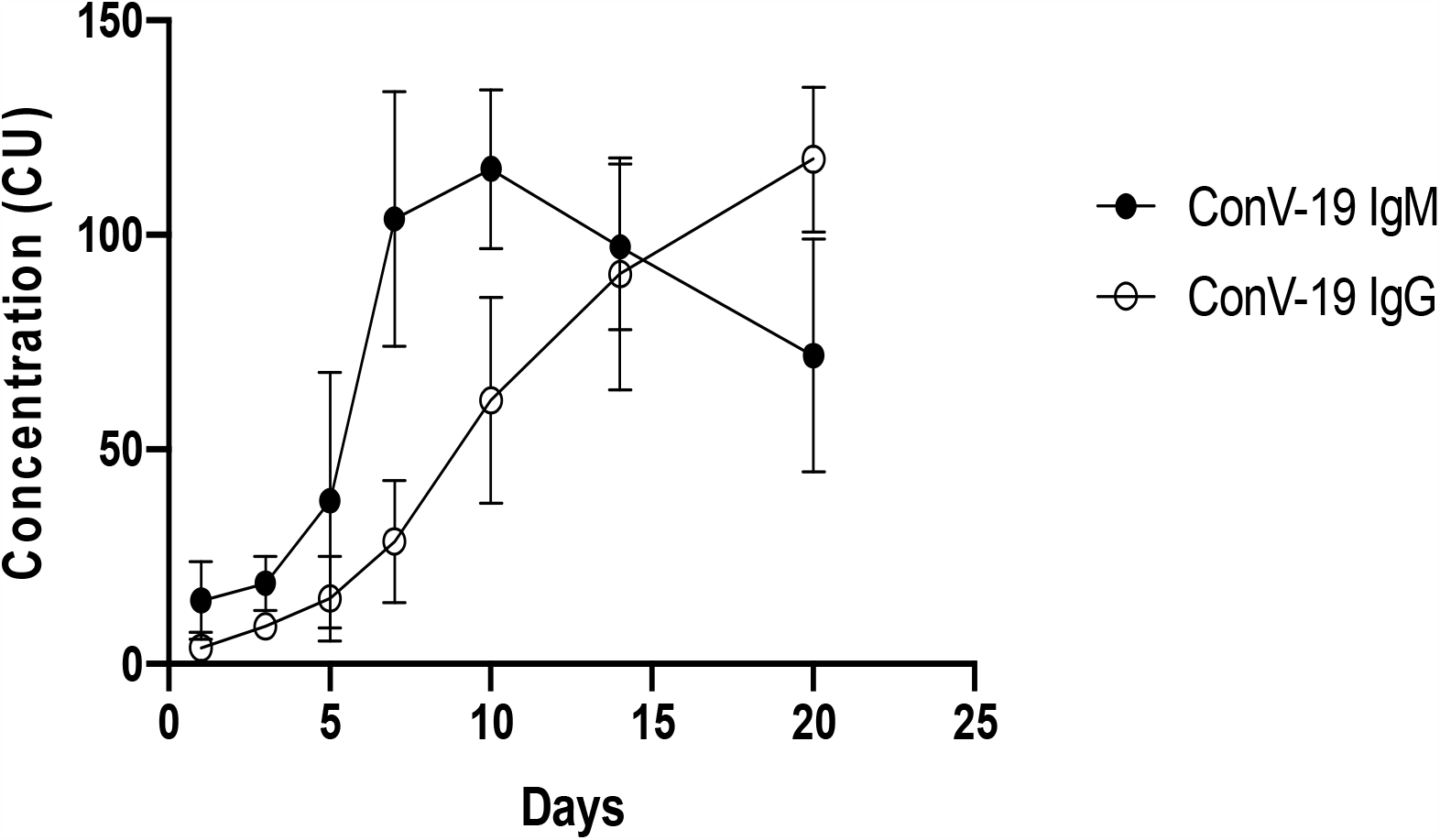
The change pattern of antibodies against SARS-CoV-2 of COVID-19 disease in continuous monitoring patients.

## Discussion

In this study, we investigated the overall profile and kinetics of the COVID-19 antibody response in four regions of China, including Wuhan, Beijing, Tianjin, Shijiazhuang, of which Wuhan is a high incidence area of SARS-CoV-2 infection disease in early 2020[5]. In the cross-sectional study, 91.9% of 235 patients confirmed SARS-CoV-2 infection tested IgG positive and 92.3% IgM positive, accordingly 2.1% of 336 suspected patients tested IgG postitive and 5.4% IgM postive. 30 patients diagnosed with SARS-CoV-2 infection were enrolled in the cohort study for flowing-up in 20 days. The peak of IgM and IgG reached in 10^th^ and 20 ^th^ day separately after symptom onset. The positive rate of COVID-19 IgG and IgM increased along with the clinical classification and the delay of treatment time.

Facing to the worldwide prevalence of COVID-19 disease, urgent demand has spawned a large number of in vitro diagnostic products. The range of sesitivity and specificity of these products was reported as 88-100% and 75-100%[6-8]^(Shaw Andrew M,Hyde Christopher,Merrick Blair et al. Real-world evaluation of a novel technology for quantitative simultaneous antibody detection against multiple SARS-CoV-2 antigens in a cohort of patients presenting with COVID-19 syndrome.[J]. Analyst, 2020, undefined: undefined.)(GeurtsvanKessel Corine H,Okba Nisreen M A,Igloi Zsofia et al. An evaluation of COVID-19 serological assays informs future diagnostics and exposure assessment.[J] .Nat Commun, 2020, 11: 3436.).^ In our study, approximately 8% COVID-19 confirmed patients were negative for IgM or IgG, respectively. Possible reasons might lead to negative results for confirmed patients. Firstly, various detection time points of patients were an important factor[10,11], since it was reported that the antibody responses to SARS-CoV-2 are three days later from symptom onset or one week later after infection with SARS-CoV-2[12]. In our study Figure 1A also shows antibody positive rate increased along with the symptom onset intervals, the seroprevalence of IgM and IgG were 81% and 83% less than day 7 after onset of symptoms and reached over 95% in two weeks later, a little higher than a previous study[9]. Therefore, the production of specific antibody exists window phase after individuals were infected with SARS-CoV-2. COVID-19 patients tested antibody in window phase also could lead to false-negative result due to the humoral immunity not respond during the early infection period[13,14]. Secondly, COVID-19 patients may not produce the specific antibody test against SARS-CoV-2 in essence. Some COVID-19 patients were negative for IgM and IgG from onset to recovery[15], which indicated that innate immune could make the virus clearance without the function of adaptive immune[14,15]. We found that the seropositive rates for IgM and IgG antibody in COVID-19 patients from Shijiazhuang and Tianjin were higher than COVID-19 patients from Beijing and Wuhan. The varied seropositive rates in different regions were likely associated with the detected time point of antibody tests which may affect the false negative results. Additionally, the sample size for COVID-19 patients are less in Beijing and Wuhan, which also may lead to the seropositive rates for IgM and IgG antibody are lower[16]. For non-infected patients there’s also 2.1% for IgG postitivity and 5.4% IgM postivitity. There’s many reasons for the false positive results, including immune disease, cancer, drug usage and other infection, etc[17,18]. So we agree it’s not a reliable tests for diagnosis of COVID-19.

The antibody titer in different clinical stages was explored in our study. IgM and IgG antibody titer in severe cases was observed higher than non-severe cases in this study, especially IgG antibody titer is especially higher in heavy cases, howerver, there was no statistically significant for the small number of cases. Our results were consistent with previous findings. Long *et al*. has reported that severe cases with higher titer of IgM and IgG antibody compared with non-severe cases[19]. Meanwhile, Qu *et al*. showed that the critical COVID-19 patients with more intensive IgM and IgG antibody responses compared with non-critical patients[20]. The two studies indicated that IgM and IgG antibody titer may be associated with the disease severity of COVID-19 patients, which could be explained by a high level of virus load or inflammatory storm in serve or critical cases[21]. Ages of severe cases were older than non-severe cases, but the difference was no statistical significance. Of note, many studies have reported that non-survivors were older than survivors in critical cases of COVID-19[22-24], especially the patients combined with basic diseases, including hypertension, coronary artery disease and diabetes[24,25]. Elder age was one of the risk factors for infected with SARS-CoV-2 and more easily progressed to critical cases with poorer prognosis[26]. Further study with more cases need to be organized to confirm our results.

The kinetics of IgM and IgG antibody responses in COVID-19 patients contribute to understand the immune responses caused by SARS-CoV-2. Figure 2 shows that the IgM antibody titer reached a peak earlier than the IgG antibody. Limited to the follow-up period, the peak titer of IgG antibody was not observed. Andrea *et al*. reported that IgM antibody levels peaked at 10–12 days and significantly declined after 18 days[27], similiar with our study. It was reported that IgG antibody usually appeared in the late stage of infection and it can persist beyond 7 weeks[10]. Although antibody test didn’t mean that COVID-19 patients have produced protective antibody against SARS-CoV-2, due to many detected antibodies were not neutralizing antibodies[14,15,28]. Some studies has shown COVID-19 patients with high titer of IgG might have produced neutralizing antibody which can make virus clearance[29,30]. Therefore, the results of COVID-19 antibody test should be explained with caution when used for evaluating the patient’s condition.

We conducted a multi-center study and a cohort study to investigate the change characters of IgM and IgG in the progress of COVID-19 disease to better understand the immune response in COVID-19 patients after they infected with SARS-CoV-2. However, further research were needed for more cases enrolled. Additionally, individuals with asymptomatic SARS-CoV-2 infection were not included in our studies, so an epidemiological investigation needed to conduct.

## Data Availability

The data used to support the findings of this study are available from the corresponding author upon request.

## Acknowledgments

The manufacturer provided equipment and reagent for this study, but had no role in directing the study or influencing the study outcomes.

## Author contributions

All authors have accepted responsibility for the entire content of this manuscript and approved its submission.

## Informed consent

Informed consent was waived for remaining samples left after measurement in this study.

## Ethical approval

The study has been cleared by the ethnic committee of Peking Union Medical College Hospital (ZS-2303).

## References

1. NHC (2020) Chinese Recommendations for Diagnosis and Treatment of SARS-CoV-2 infection. http://www.nhc.gov.cn/xcs/zhengcwj/202003/46c9294a7dfe4cef80dc7f5912eb1989.shtml. Accessed 07.07 2020

2. To KK, Tsang OT, Chik-Yan Yip C, Chan KH, Wu TC, Chan JMC, Leung WS, Chik TS, Choi CY, Kandamby DH, Lung DC, Tam AR, Poon RW, Fung AY, Hung IF, Cheng VC, Chan JF, Yuen KY (2020) Consistent detection of 2019 novel coronavirus in saliva. Clinical infectious diseases : an official publication of the Infectious Diseases Society of America. doi:10.1093/cid/ciaa149

3. Pan Y, Zhang D, Yang P, Poon LLM, Wang Q (2020) Viral load of SARS-CoV-2 in clinical samples. The Lancet Infectious diseases 20 (4):411–412. doi:10.1016/s1473-3099(20)30113-4

4. Lerner AM, Eisinger RW, Lowy DR, Petersen LR, Humes R, Hepburn M, Cassetti MC (2020) The COVID-19 Serology Studies Workshop: Recommendations and Challenges. Immunity. doi:10.1016/j.immuni.2020.06.012

5. Wu JT, Leung K, Leung GM (2020) Nowcasting and forecasting the potential domestic and international spread of the 2019-nCoV outbreak originating in Wuhan, China: a modelling study. Lancet (London, England) 395 (10225):689–697. doi:10.1016/s0140-6736(20)30260-9

6. Haselmann V, Kittel M, Gerhards C, Thiaucourt M, Eichner R, Costina V, Neumaier M (2020) Comparison of test performance of commercial anti-SARS-CoV-2 immunoassays in serum and plasma samples. Clinica chimica acta; international journal of clinical chemistry 510:73–78. doi:10.1016/j.cca.2020.07.007

7. Shaw AM, Hyde C, Merrick B, James-Pemberton P, Squires BK, Olkhov RV, Batra R, Patel A, Bisnauthsing K, Nebbia G, MacMahon E, Douthwaite S, Malim M, Neil S, Martinez Nunez R, Doores K, Mark TKI, Signell AW, Betancor G, Wilson HD, Galão Rp, Pickering S, Edgeworth JD (2020) Real-world evaluation of a novel technology for quantitative simultaneous antibody detection against multiple SARS-CoV-2 antigens in a cohort of patients presenting with COVID-19 syndrome. The Analyst. doi:10.1039/d0an01066a

8. GeurtsvanKessel CH, Okba NMA, Igloi Z, Bogers S, Embregts CWE, Laksono BM, Leijten L, Rokx C, Rijnders B, Rahamat-Langendoen J, van den Akker Jpc, van Kampen Jja, van der Eijk AA, van Binnendijk RS, Haagmans B, Koopmans M (2020) An evaluation of COVID-19 serological assays informs future diagnostics and exposure assessment. Nature communications 11 (1):3436. doi:10.1038/s41467-020-17317-y

9. Xu X, Sun J, Nie S, Li H, Kong Y, Liang M, Hou J, Huang X, Li D, Ma T, Peng J, Gao S, Shao Y, Zhu H, Lau JY, Wang G, Xie C, Jiang L, Huang A, Yang Z, Zhang K, Hou FF (2020) Seroprevalence of immunoglobulin M and G antibodies against SARS-CoV-2 in China. Nature medicine. doi:10.1038/s41591-020-0949-6

10. Sethuraman N, Jeremiah SS, Ryo A (2020) Interpreting Diagnostic Tests for SARS-CoV-2. Jama. doi:10.1001/jama.2020.8259

11. Zainol Rashid Z, Othman SN, Abdul Samat MN, Ali UK, Wong KK (2020) Diagnostic performance of COVID-19 serology assays. The Malaysian journal of pathology 42 (1):13–21

12. Lauer SA, Grantz KH, Bi Q, Jones FK, Zheng Q, Meredith HR, Azman AS, Reich NG, Lessler J (2020) The Incubation Period of Coronavirus Disease 2019 (COVID-19) From Publicly Reported Confirmed Cases: Estimation and Application. Annals of internal medicine 172 (9):577–582. doi:10.7326/m20-0504

13. Lou B, Li TD, Zheng SF, Su YY, Li ZY, Liu W, Yu F, Ge SX, Zou QD, Yuan Q, Lin S, Hong CM, Yao XY, Zhang XJ, Wu DH, Zhou GL, Hou WH, Li TT, Zhang YL, Zhang SY, Fan J, Zhang J, Xia NS, Chen Y (2020) Serology characteristics of SARS-CoV-2 infection since exposure and post symptom onset. The European respiratory journal. doi:10.1183/13993003.00763-2020

14. Younes N, Al-Sadeq DW, Al-Jighefee H, Younes S, Al-Jamal O, Daas HI, Yassine HM, Nasrallah GK (2020) Challenges in Laboratory Diagnosis of the Novel Coronavirus SARS-CoV-2. Viruses 12 (6). doi:10.3390/v12060582

15. Wang B, Wang L, Kong X, Geng J, Xiao D, Ma C, Jiang XM, Wang PH (2020) Long-term coexistence of SARS-CoV-2 with antibody response in COVID-19 patients. Journal of medical virology. doi:10.1002/jmv.25946

16. Freiman JA, Chalmers TC, Smith H, Jr., Kuebler RR (1978) The importance of beta, the type II error and sample size in the design and interpretation of the randomized control trial. Survey of 71 “negative” trials. The New England journal of medicine 299 (13):690–694. doi:10.1056/nejm197809282991304

17. Wang Q, Du Q, Guo B, Mu D, Lu X, Ma Q, Guo Y, Fang L, Zhang B, Zhang G, Guo X (2020) A Method To Prevent SARS-CoV-2 IgM False Positives in Gold Immunochromatography and Enzyme-Linked Immunosorbent Assays. Journal of clinical microbiology 58 (6). doi:10.1128/jcm.00375-20

18. Deeks JJ, Dinnes J, Takwoingi Y, Davenport C, Spijker R, Taylor-Phillips S, Adriano A, Beese S, Dretzke J, Ferrante di Ruffano L, Harris IM, Price MJ, Dittrich S, Emperador D, Hooft L, Leeflang MM, Van den Bruel A (2020) Antibody tests for identification of current and past infection with SARS-CoV-2. The Cochrane database of systematic reviews 6:Cd013652. doi:10.1002/14651858.Cd013652

19. Long QX, Liu BZ, Deng HJ, Wu GC, Deng K, Chen YK, Liao P, Qiu JF, Lin Y, Cai XF, Wang DQ, Hu Y, Ren JH, Tang N, Xu YY, Yu LH, Mo Z, Gong F, Zhang XL, Tian WG, Hu L, Zhang XX, Xiang JL, D. HX Liu HW, Lang CH, Luo XH, Wu SB, Cui XP, Zhou Z, Zhu MM, Wang J, Xue CJ, Li XF, Wang L, Li ZJ, Wang K, Niu CC, Yang QJ, Tang XJ, Zhang Y, Liu XM, Li JJ, Zhang DC, Zhang F, Liu P, Yuan J, Li Q, Hu JL, Chen J, Huang AL (2020) Antibody responses to SARS-CoV-2 in patients with COVID-19. Nature medicine 26 (6):845–848. doi:10.1038/s41591-020-0897-1

20. Qu J, Wu C, Li X, Zhang G, Jiang Z, Li X, Zhu Q, Liu L (2020) Profile of IgG and IgM antibodies against severe acute respiratory syndrome coronavirus 2 (SARS-CoV-2). Clinical infectious diseases : an official publication of the Infectious Diseases Society of America. doi:10.1093/cid/ciaa489

21. Yongchen Z, Shen H, Wang X, Shi X, Li Y, Yan J, Chen Y, Gu B (2020) Different longitudinal patterns of nucleic acid and serology testing results based on disease severity of COVID-19 patients. Emerging microbes & infections 9 (1):833–836. doi:10.1080/22221751.2020.1756699

22. Chen N, Zhou M, Dong X, Qu J, Gong F, Han Y, Qiu Y, Wang J, Liu Y, Wei Y, Xia J, Yu T, Zhang X, Zhang L (2020) Epidemiological and clinical characteristics of 99 cases of 2019 novel coronavirus pneumonia in Wuhan, China: a descriptive study. Lancet (London, England) 395 (10223):507–513. doi:10.1016/s0140-6736(20)30211-7

23. Yang X, Yu Y, Xu J, Shu H, Xia J, Liu H, Wu Y, Zhang L, Yu Z, Fang M, Yu T, Wang Y, Pan S, Zou X, Yuan S, Shang Y (2020) Clinical course and outcomes of critically ill patients with SARS-CoV-2 pneumonia in Wuhan, China: a single-centered, retrospective, observational study. The Lancet Respiratory medicine 8 (5):475–481. doi:10.1016/s2213-2600(20)30079-5

24. Zhou F, Yu T, Du R, Fan G, Liu Y, Liu Z, Xiang J, Wang Y, Song B, Gu X, Guan L, Wei Y, Li H, Wu X, Xu J, Tu S, Zhang Y, Chen H, Cao B (2020) Clinical course and risk factors for mortality of adult inpatients with COVID-19 in Wuhan, China: a retrospective cohort study. Lancet (London, England) 395 (10229):1054–1062. doi:10.1016/s0140-6736(20)30566-3

25. Zhang P, Zhu L, Cai J, Lei F, Qin JJ, Xie J, Liu YM, Zhao YC, Huang X, Lin L, Xia M, Chen MM, Cheng X, Zhang X, Guo D, Peng Y, Ji YX, Chen J, She ZG, Wang Y, Xu Q, Tan R, Wang H, Lin J, Luo P, Fu S, Cai H, Ye P, Xiao B, Mao W, Liu L, Yan Y, Liu M, Chen M, Zhang XJ, Wang X, Touyz RM, Xia J, Zhang BH, Huang X, Yuan Y, Rohit L, Liu PP, Li H (2020) Association of Inpatient Use of Angiotensin-Converting Enzyme Inhibitors and Angiotensin II Receptor Blockers With Mortality Among Patients With Hypertension Hospitalized With COVID-19. Circulation research 126 (12):1671–1681. doi:10.1161/circresaha.120.317134

26. Davies NG, Klepac P, Liu Y, Prem K, Jit M, Eggo RM (2020) Age-dependent effects in the transmission and control of COVID-19 epidemics. Nature medicine. doi:10.1038/s41591-020-0962-9

27. Padoan A, Sciacovelli L, Basso D, Negrini D, Zuin S, Cosma C, Faggian D, Matricardi P, Plebani M (2020) IgA-Ab response to spike glycoprotein of SARS-CoV-2 in patients with COVID-19: A longitudinal study. Clinica chimica acta; international journal of clinical chemistry 507:164–166. doi:10.1016/j.cca.2020.04.026

28. Yu F, Du L, Ojcius DM, Pan C, Jiang S (2020) Measures for diagnosing and treating infections by a novel coronavirus responsible for a pneumonia outbreak originating in Wuhan, China. Microbes and infection 22 (2):74–79. doi:10.1016/j.micinf.2020.01.003

29. Perera RA, Mok CK, Tsang OT, Lv H, Ko RL, Wu NC, Yuan M, Leung WS, Chan JM, Chik TS, Choi CY, Leung K, Chan KH, Chan KC, Li KC, Wu JT, Wilson IA, Monto AS, Poon LL, Peiris M (2020) Serological assays for severe acute respiratory syndrome coronavirus 2 (SARS-CoV-2), March 2020. Euro surveillance : bulletin Europeen sur les maladies transmissibles = European communicable disease bulletin 25 (16). doi:10.2807/1560-7917.Es.2020.25.16.2000421

30. To KK, Tsang OT, Leung WS, Tam AR, Wu TC, Lung DC, Yip CC, Cai JP, Chan JM, Chik TS, Lau DP, Choi CY, Chen LL, Chan WM, Chan KH, Ip JD, Ng AC, Poon RW, Luo CT, Cheng VC, Chan JF, Hung IF, Chen Z, Chen H, Yuen KY (2020) Temporal profiles of viral load in posterior oropharyngeal saliva samples and serum antibody responses during infection by SARS-CoV-2: an observational cohort study. The Lancet Infectious diseases 20 (5):565–574. doi:10.1016/s1473-3099(20)30196-1

